# FAMILY’S RESILIENCE IN CAREGIVING ELDERLY WITH DEMENTIA: A SYSTEMATIC REVIEW

**DOI:** 10.1101/2021.06.16.21259058

**Authors:** Anung Ahadi Pradana, Rohayati

## Abstract

**Objective:** Dementia patients have complex needs that may become a burden to those who caregive them. They are treated at home by family members due to financial limitations and lack of support from health services. Therefore, the measurement of self-resilience in caregivers needs special attention from professionals.

**Method:** This study used a systematic review method from 4 databases consisting of the CINAHL, ProQuest, PubMed, and Google Scholar between 2015 till 2020; 17 articles were considered to meet the inclusion criteria.

**Results:** Caregiver of elderly with dementia has a tendency to experience burdens that affect the quality of the care performed. The level of resilience that caregivers have can help reduce the level of burden they experience and improve their quality of life.

**Conclusions:** Social support and formal support by health workers for caregivers have a significantly positive effect in increasing the level of resilience of caregivers.

## Introduction

Dementia has become a global disease that affects around 50 million people with 7.7 million new patients every year in the world, with 60% of whom live in developing countries. The number of dementia patients is estimated to increase to 82 million in 2030 and 152 million in 2050.^1^ Alzheimer Indonesia^2^ states that the number of dementia patients in Indonesia in 2010 were 1.2 million people, while it is estimated that in 2020 it will be 2 million and become 4 million in 2050.

Most of dementia patients are treated at home by family members due to financial limitations and lack of support from health services.^2^ The full dependence needed by dementia patients can sometimes be a trigger for physical and emotional exhaustion from family members. Behavioral problems and psychological symptoms that occur in dementia patients as well as socio-demographic factors and psychological factors from the family of caregivers are 3 main factors that become challenges in providing care by families.^3^ The burden experienced by the caregiver can be influenced by the level of self-resilience.^4^

Resilience is a process in which individuals exhibit positive adaptation despite exposure to adverse life events, such as a diagnosis of dementia.^4^ These subjective measures can be biased by people’s tendency to overestimate their resilient abilities or underestimate it in response to conditions such as depression. Caregivers who consider themselves highly resilient are often not tough and vice versa.^5^ Therefore, the measurement of self-resilience in caregivers needs special attention from professionals.

## Method

This study was a systematic review. The following databases were searched in July 2020: Cumulative Index to Nursing and Allied Health Literature (CINAHL), ProQuest, PubMed, and Google Scholar between 2015 till 2020. The keywords used were a combination of “resilience”, “elderly”, “family”, and “caregiver”. Studies were included if (1) articles in English, (2) qualitative/quantitative research, (3) articles about the family’s resilience on caregiving older adults with dementia. Exclusion criteria consisted of (1) articles in another language, (2) articles conducted in review article, (3) main caregiver is not family.

## Results

From a total of 127 articles, 17 articles were considered to meet the inclusion criteria and passed the screening process using the PRISMA diagram (Diagram 1). Based on 17 journal articles obtained, 8 articles used qualitative methods, 8 articles used cross-sectional quantitative methods, and 1 other article used mixed methods. The critical review that was carried out followed the Critical Appraisal Skills Program (CASP) guidelines from Cochrane, 17 selected articles were analyzed using the RobVis’s Tracking Media to obtain the Risk of Bias extraction results as listed in Figure 2.

**Figure 1.**
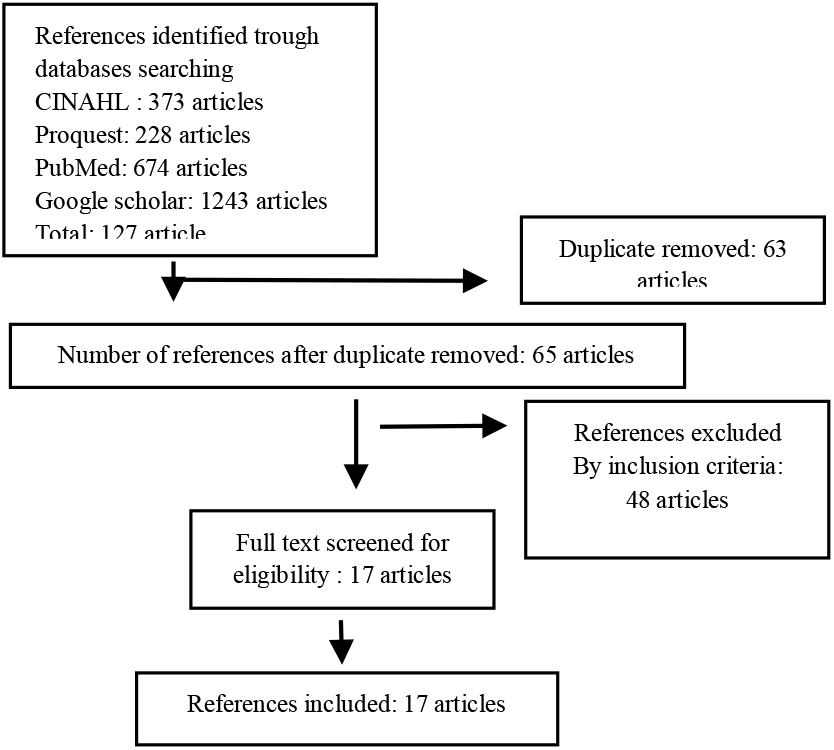
PRISMA Diagram.

**Figure 2.**
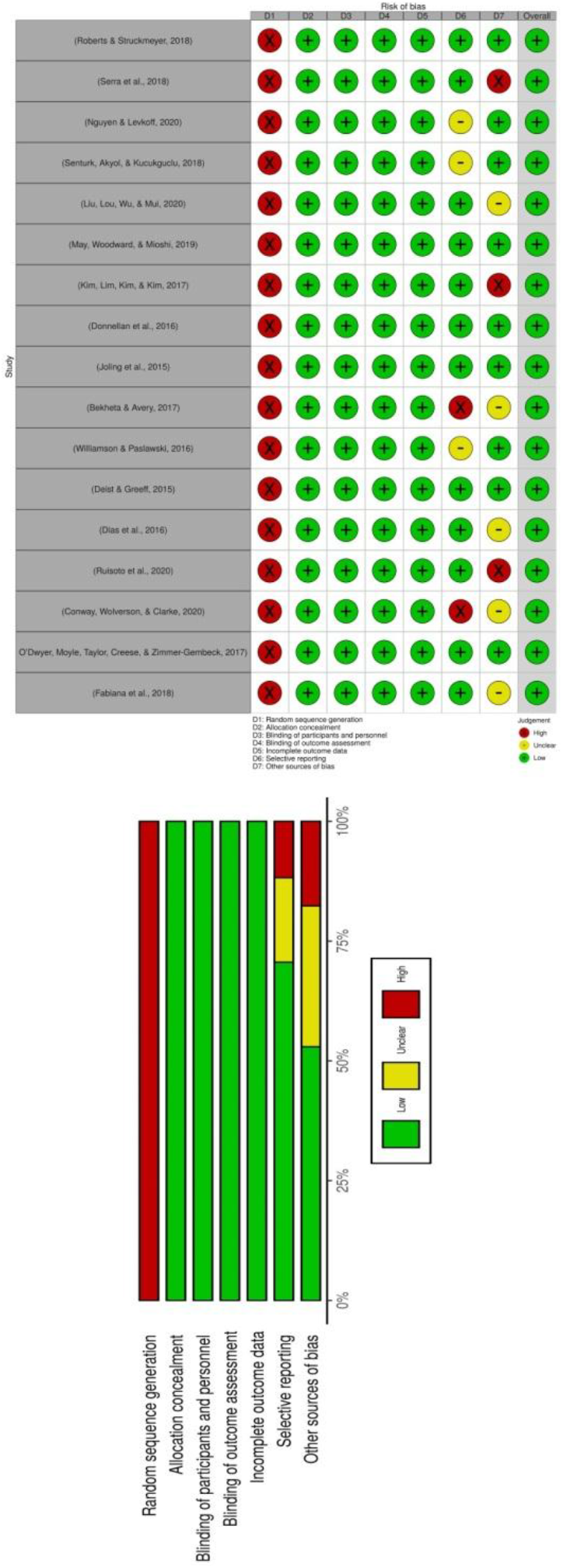
Risk of Bias.

## Discussions

Caregivers for older adults with dementia experience a relatively high burden, especially in the face of family dynamics, isolation, financial struggles, seeking respect, and acceptance.^6^ Research shows that the severity of dementia level condition experienced by elderly is directly proportional to the burden of caregiving experienced by the family. This is exacerbated by an increase in caregiving hours, a bad relationship between caregivers and elderly.^7^ The increased burden of caregivers experienced can increase the risk of abuse in elderly.

Caregiver’s burden can be reduced by increasing the level of resilience of the caregiver which can be done by providing social support and formal support for the caregiver, and helping the caregiver develop effective coping techniques.^8^ Caregiver’s resilience can improve family adaptation patterns to dementia problems experienced by elderly.^9^ The level of caregivers’ resilience is influenced by several factors including internal and external factors. Internal factors include demographic factors from the family, personal beliefs, caregiving commitment, and the caregivers’ coping ability in dealing with adversity. Meanwhile, external factors include cultural background, belief systems, family economic status, communication processes in the family, stress experienced by the family, social support, problematic behavior of elderly with dementia, and pressure from the community.^10-12^

Research shows that the social support things are often viewed differently by caregivers. Caregivers consider that support from family is not very influential because the assistance provided is relatively low compared to support from friends, especially those who also experience the same thing. The available social support should focus on internal/external factors that can increase resilience to caregivers.^13^

Resilient condition is a subjective response experienced by each caregiver in facing adversity that appears. Research conducted by Bekheta & Avery^14^ shows that families perceive that resilience is identified as the ability to survive from risk factors to being carers, which include stress, frustration, lack of social support, fatigue, and negative feelings (sadness, anger). Other factors related to the caregiver’s view of resilience include being considered a life goal, perspectives, and resources for caregivers.^4^

For spouses who are carers, resilience is often described as a continuous process that helps them maintain couplehood, a sense of togetherness, and reciprocity in their relationship.^15^ Many caregivers consider resilient conditions as a value, a resource or a result of being caregivers.^16^ Other studies have shown that resilient caregivers have a high quality of life, low depressive symptoms, and are able to care for elderly with relatively severe dementia conditions.^17^

## Conclusions

Caregiver of elderly with dementia has a tendency to experience burdens that affect the quality of care performed. The level of resilience that caregivers have can help reduce the level of burden they experience and improve their quality of life. Social support and formal support by health workers for caregivers have a significantly positive effect in increasing the level of resilience of caregivers.

## Data Availability

Tee Data Availability is open to access

## Acknowledgements

This work is supported and funded by STIKes Mitra Keluarga.

## Notes

### Competing Interest Statement

The authors have declared no competing interest.

### Author Declarations

All authors have assented to posting of the manuscript and inclusion as authors.

